# Quantification of Epicardial Fat Volume as a Novel Cardiovascular Risk Marker in Asymptomatic Subjects for Early Detection of Cardiovascular Disease

**DOI:** 10.1101/2020.06.28.20141622

**Authors:** Mahfouz El Shahawy, Lillee Izadi, Antonella Sabatini, Susan Tucker, Jessica Patella, Omar El Shahawy, Frederick Yturralde

## Abstract

Epicardial fat volume (EFV), also known as epicardial adipose tissue (EAT), sometimes acts as a protector against heart problems; however, in excess volume was found to be associated with cardiovascular structural and functional abnormalities. This study aims to establish a threshold between normal and abnormal values for EFV/EAT in asymptomatic subjects, as well as to assess whether excess EFV/EAT is associated with significant structural and functional abnormalities, including coronary artery calcium score (CACS). A total of 220 asymptomatic patients, were screened utilizing Early Cardiovascular Disease Risk Score (ECVDRS), and CT for CACS and EFV/EAT quantification. Out of the 220 subjects, 69 had a 0 CACS and were included in this analysis. These 69 were then further categorized into 3 groups: Group 1 (Normal subjects; n=20) with ECVDRS < 3, and ACC/AHA risk score < 5%; Group 2 (n= 16) with elevated EFV/EAT and no abdominal visceral obesity; Group 3 (n=33) with elevated EFV/EAT and abdominal visceral obesity. The average EFV/EAT was identified to be 69 cm^3^ ± 20 in females and 68 cm^3^ ± 15 in males among Group 1, which indicate the normal values for EFV/EAT. It was also found that elevated EFV/EAT without (Group 2) or with (Group 3) abdominal visceral adiposity was associated with significant vascular abnormalities, as compared to the normal group among these populations of asymptomatic patients with 0 CACS. Elevated EFV/EAT is a novel cardiovascular risk marker regardless of gender, which might be the culprit for major cardiovascular risk factors.

## Introduction

Epicardial fat volume (EFV), also known as epicardial adipose tissue (EAT), has been recently reported to be a potential novel cardiovascular risk marker.^1-5^ Early detection of an abnormal EFV/EAT (i.e. > 90cm^3^ ± 20 in females or males) has been found to correlate with early cardiovascular disease among subjects with type 2 diabetes and obesity, which mandates early treatments, including aggressive lifestyle modifications and aggressive medical therapy.^6^ EFV/EAT has different mRNA than that of abdominal visceral adipose tissue. Epicardial fat belongs to the category of perivascular adipose tissue which also includes the renal arteries. It has been reported that many obese and/or diabetic individuals have higher EFV/EAT, which has been associated with coronary artery disease (CAD) risk factors.^2,3,7^ Additionally, other studies found an association between increased accumulations of EFV/EAT and metabolic syndrome.^8,9^ EFV/EAT has also been associated with multiple independent cardiac risk factors, such as high fasting glucose, high levels of C-reactive protein, low HDL and carotid intima-media thickness.^10^

### Epicardial fat structure and functionality

Epicardial fat (EF) is defined as perivascular adipose tissue which deposits over the heart, and it can cover 80% of the heart muscle while making up 20% of the heart’s mass.^5,11,12^ Epicardial fat is vascularized by branches of the coronary arteries, and has no fascia layer separating it from the myocardium, hence it shares the same microcirculation, suggesting a close and strong interaction with both tissue structures.^12^ The physiologic function of EF is complex and not yet completely understood or elucidated.^11,12^ Its function can be generally distinguished by mechanical, metabolic, thermogenic, and endocrine/paracrine functions.^12^ A higher rate of free fatty acid release and uptake from EF compared to subcutaneous and other visceral fat depots suggests that it has a role in sustaining myocardium energy supplies.^13^ Additionally, there is growing evidence that human EF produces bioactive cytokines, known as secretomes. These cytokines are involved in the regulation of endothelial function, where EF has been shown to be the strongest predictor of endothelial dysfunction via abnormal local pulse wave velocity in carotid arterial stiffness in menopausal women ^14^. Secretomes also regulate coagulation and inflammation.^12^ In fact, several bioactive molecules secreted from EF can either protect or adversely affect the myocardium and the coronary arteries.^12^ Under normal physiological conditions, fat produces anti-inflammation or anti-atherosclerotic cytokines, such as adiponectin and adrenomedullin, to exert its cardioprotective functions.^15,16^ Adiponectin has been described as having anti-diabetic, anti-atherogenic, anti-oxidative and anti-inflammatory properties, and is only produced by adipocytes.^16^ It activates the adenosine monophosphate-activated protein kinase (AMPK) pathway, leading to increased fatty acid oxidation and reduced lipid deposits in cardiac muscle.^16^ Adenomedullin is produced in a variety of organs and exerts many cardioprotective actions which include vasodilation, natriuresis, anti-apoptosis, and stimulation of nitric oxide production.^17^

### Pathophysiology of excessive Epicardial fat volume

Excessive accumulation of EF is associated with cardiovascular structural and functional abnormalities.^1-3,7^ It can lead to additional mass on both ventricles that can increase the work demand of the heart, resulting in left ventricular hypertrophy.^18^ The first evidence that implicated the inflammatory state of human EF was in 2003 by Mazurek, et al.^19^ Since then, many studies have shown elevated inflammation with EF in terms of leukocyte infiltration and inflammatory gene expression.^12,15^ Macrophages are one of the major effector cells mediating adipose inflammation.^12^ It has been shown that patients with advanced coronary artery disease (CAD) have more inflammatory M1 (classically activated) macrophages but free anti-inflammatory M2 (alternatively activated) macrophages in EF compared to subjects without CAD.^20^ There are additional complex interactions with T lymphocytes involving Toll-like receptors (TLRs) that lead to transcription of inflammatory markers such as interleukin-1 (IL-1), IL-6, tumor necrosis factor alpha, and monocyte chemoattractant protein-1.^21^ Thus, the inflammatory process within EF results in paracrine or vasocrine secretion of epicardial inflammatory cytokines, in turn creating the metabolic and inflammatory milieu that promotes CAD.

Early detection of cardiovascular disease is of the utmost importance. While using the Early Cardiovascular Disease Risk Score (ECVDRS) over the past decade, many of our prior publications underscored the gap of identifying early cardiovascular risk markers.^7^ The current study attempts to address this gap by exploring the potential of EFV/EAT normal value determination, in addition to ECVDRS among subjects with 0 CACS score, will lead to more accurate assessment of cardiovascular risks. This may allow clinicians to offer early lifestyle interventions to possibly delay, or even prevent, the *Atherosclerotic Cardiovascular Disease (*ASCVD).

## Methods

We screened 2,756 asymptomatic subjects, ages 20-79, for CVD risk using the Early Cardiovascular Disease Risk Scoring System (ECVDRS), which consists of 10 tests; 7 of these tests are vascular and 3 are cardiac. The vascular tests are as follows: large (C1) and small (C2) artery stiffness, blood pressure (BP) at rest and post-mild protocol exercise (PME), Carotid Intima Media Thickness (CIMT), abdominal aorta ultrasound, retinal photography, and microalbuminuria. The 3 cardiac tests are as follows: Pro-BNP, ECG, and left ventricle ultrasound (LVUS). Additional tests are waist circumference, BMI, fasting blood sugar, lipid profile, and hs-CRP. The current ACC/AHA risk score was also calculated to assess and estimate the 10-year risk for the development of ASCVD. Out of the total, 596 subjects were asymptomatic, and out of these subjects, 220 (37%) underwent cardiac CT for CACS and EFV determination using Siemens Somatom Definition Dual source CT scanner 64×2. A total of 69 out of the 220 subjects had a 0 CACS. These 69 subjects were divided into 3 groups: Group 1: 20 normal subjects with the following metrics: ECVDRS < 3, ACC/AHA risk score < 5%, CACS 0. Group 2: 16 subjects who had elevated EFV and no abdominal visceral obesity. Group 3: 33 subjects who had elevated EFV and abdominal visceral adiposity.

## Results

As shown in Table 1, normal EFV/EAT in Group 1 was determined to be 69 cm^3^ ± 20 in females and 68 cm^3^ ± 15 in males and was associated with minimal structural and functional abnormalities. Group 2 increased EFV/EAT values were associated with statistically significant structural and functional abnormalities regardless of sex, as compared with Group 1. Group 3 increased EFV/EAT with abdominal visceral adiposity was associated with increases in structural and functional vascular abnormalities that were statistically significant compared to Group 1, but not significant in relation to Group 2, implying that EFV/EAT is the primary culprit for these vascular abnormalities, particularly an abnormal rise in BP-PME (p < 0.0001).

**Table 1:** Excess EFV/EAT is associated with CV structural and functional abnormalities

|  | Group 1 |  | Group 2 |  | Group 3 |  | P-values |
| --- | --- | --- | --- | --- | --- | --- | --- |
|  | Female | Male | Female | Male | Female | Male |  |
|  | 14 (67%) | 6 (33%) | 10 (71%) | 6 (29%) | 16 (48%) | 17 (52%) |  |
| **EFV Average ($\text{cm}^3$)** | $69 \pm 20$ | $68 \pm 15$ | $106 \pm 18$ | $114 \pm 20$ | $125 \pm 20$ | $160 \pm 32$ | < 0.004 |
| **ECVDRS Avg** | 3.07 | 1.5 | 6.70 | 5.50 | 6.81 | 5.88 | < 0.014 |
| **ECVDRS Total Score** | 0-3 | 0-3 | 4-10 | 2-9 | 1-16 | 0-13 |  |
| **Vascular Avg** | 2.23 | 1.3 | 7.00 | 4.66 | 5.56 | 4.58 | < 0.0001 |
| **Cardiac Avg** | 0.28 | 0.16 | 1.00 | 0.5 | 1.25 | 1.24 | < 0.01 |
| **AHA/ACC Risk Score (%) Avg** | 1.79 | 3.73 | 3.66 | 8.73 | 6.49 | 10.82 | < 0.15 |
| **Abnormal BP rise in PME** | 5 (36%) | 2 (29%) | 8 (80%) | 5 (83%) | 11 (69%) | 12 (70%) | < 0.05 |
| **Abnormal CIMT** | 7 (50%) | 1 (7%) | 5 (50%) | 2 (33%) | 7 (44%) | 5 (29%) | < 1.00 |
| **Abnormal C1** | 1 (7%) | 0 (0%) | 6 (60%) | 1 (17%) | 9 (56%) | 2 (18%) | < 0.0001 |
| **Abnormal C2** | 5 (36%) | 0 (0%) | 6 (60%) | 4 (67%) | 8 (50%) | 8 (47%) | < 0.07 |

## Discussion

Normal EFV is associated with minimal CV structural and functional abnormalities, while excess EFV is associated with statistically significant increases in cardiovascular risk factors and comorbidities. This has been observed repeatedly over the last decade by our team and has been reported in many prior national and international cardiological congresses.^22-36^

Several measurements of fat volumes are feasible: total intrathoracic fat volume, epicardial adipose tissue and thoracic fat (i.e., ITFv, EATv, and TF volume) which can be directly measured by non-contrast cardiac computed tomography (CT).^3^ Studies have found EFV/EAT to be 22% thicker in patients ages 65 years and older, implying that EFV/EAT increases with age.^37^ There appears to be no gender difference in regards to the impact of epicardial fat volume on CV risk factors.^38^

## Conclusions

Normal EFV is associated with minimal CV structural and functional abnormalities, while excess EFV/EAT, as seen in subjects with and without adiposity, is associated with statistically significant increases in CV risk factors and comorbidities.

Given the widespread of obesity among adults and adolescents in the United States and worldwide,^39,40^ understanding the impact that EFV/EAT has on cardiovascular disease risk factors can be very important in improving cardiovascular health.

Early detection and quantification of excess EFV/EAT will help early identification and stratification of cardiovascular risk with significant guidance for early application of optimal cardiovascular treatment, including not only lifestyle modifications, but also the utilization of newer drugs which have been reported to reduce EFV/EAT.^6^ This will further prevent or delay the onset of cardiovascular disease.

As the saying goes: one ounce of early cardiovascular disease prevention is better than pounds of late treatments. *Early detect to protect; the sooner the better*.

## Limitations of the study

Only 20 subjects with 0 CACS and normal or low risk ECVDR and ACC/AHA risk scores were used as normal control. Hopefully, future studies with a greater number of subjects and stricter criteria like ours will be able to duplicate and confirm our findings.

## Data Availability

All the data is available.

## Acknowledgments

Thanks to Sarasota Memorial Health Care Foundation, Cardiovascular Center of Sarasota Foundation for Research and Education and El Shahawy Family Foundation for supporting this research. Thanks also to all subjects who participated in this study and to the many of our colleagues for referring many of their patients for CV screening. Thanks also goes particularly to cardiac surgeon Dr. Thomas Kelly, who founded the Heart Surgery Program many decades ago at our Institution, who made the comment to us that many of his patients who came for Coronary Artery Bypass Graft (CABG) had excess epicardial fat. Last but not least, thanks to our staff who has been doing an excellent job and follow up.

